# Human IgG and IgA responses to COVID-19 mRNA vaccines

**DOI:** 10.1101/2021.03.23.21254060

**Authors:** Julian Campillo-Luna, Adam V Wisnewski, Carrie A Redlich

**Affiliations:** Department of Internal Medicine, Yale University School of Medicine, New Haven, CT 06520

## Abstract

SARS-CoV-2 spike antigen-specific IgG and IgA elicited by infection mediate viral neutralization and are likely an important component of natural immunity, however, limited information exists on vaccine induced responses. We measured COVID-19 mRNA vaccine induced IgG and IgA in serum serially, up to 80 days post vaccination in 4 subjects. Spike antigen-specific IgG levels rose exponentially and plateaued 21 days after the initial vaccine dose. After the second vaccine dose IgG levels increased further, reaching a maximum approximately 7-10 days later, and remained elevated (average of 78% peak levels) during the additional 20-50 day follow up period. COVID-19 mRNA vaccination elicited spike antigen-specific IgA with similar kinetics of induction and time to peak levels, but more rapid decline in serum levels following both the 1^st^ and 2^nd^ vaccine doses (<23% peak levels within 80 days of the initial shot). The data demonstrate COVID-19 mRNA vaccines effectively induce spike antigen specific IgG and IgA and highlight marked differences in their persistence in serum.

## INTRODUCTION

Humoral responses are key components of adaptive immunity to viral infection [1]. Both alpha and gamma immunoglobulins (Ig) from COVID-19 patients mediate viral neutralization and may play distinct roles in immunity during different phases of infection and at specific anatomical sites [2-5]. IgA is the most abundantly produced Ig in humans (66 gm/kg/day) and the most abundant isotype at mucosal sites while IgG is major isotype in blood and most tissues [6, 7].

IgA’s anatomical distribution at mucosal surfaces exposed to infectious agents makes it uniquely positioned to intervene in transmission. Multiple studies have found IgA possesses superior anti-viral properties vs. IgG for influenza and for SARS-CoV-2 [8-11]. Sterlin et. al [11] recently suggested IgA dominates the early neutralizing response to SARS-CoV-2 based on multiple findings; serum IgA is 7-fold more potent than serum IgG in viral neutralization, temporal changes in circulating IgA+ plasmablasts with mucosal homing receptors, and the presence of neutralizing IgA in airway fluid and saliva.

The major SARS-CoV-2 antigenic target of human IgG and IgA is the spike protein, which is encoded by the mRNA of vaccines currently in use under EUA from the FDA [12-15]. The time course of mRNA vaccine-induced IgG responses observed during vaccine trials have recently been published [13, 14]. However, limited information exists on mRNA vaccine induced IgA responses [16]. The present study measures SARS-CoV-2 spike antigen specific serum IgA and IgG longitudinally in healthy individuals without prior COVID-19 who were among the first vaccine recipients outside of clinical trials due to their occupation as health care workers. Data from up to 80 days after the first mRNA vaccine dose are presented.

## RESULTS & DISCUSSION

Serum levels of SARS-CoV-2 spike antigen (S1 subunit)-specific IgG and IgA were measured by ELISA over time in N=4 health care workers that received COVID-19 mRNA vaccine between December 2020 and February 2021 (Table 1). The subjects had no prior history of COVID-19 and tested negative for SARS-CoV-2 nucleocapsid antigen and spike antigen at baseline.

**Table 1.**
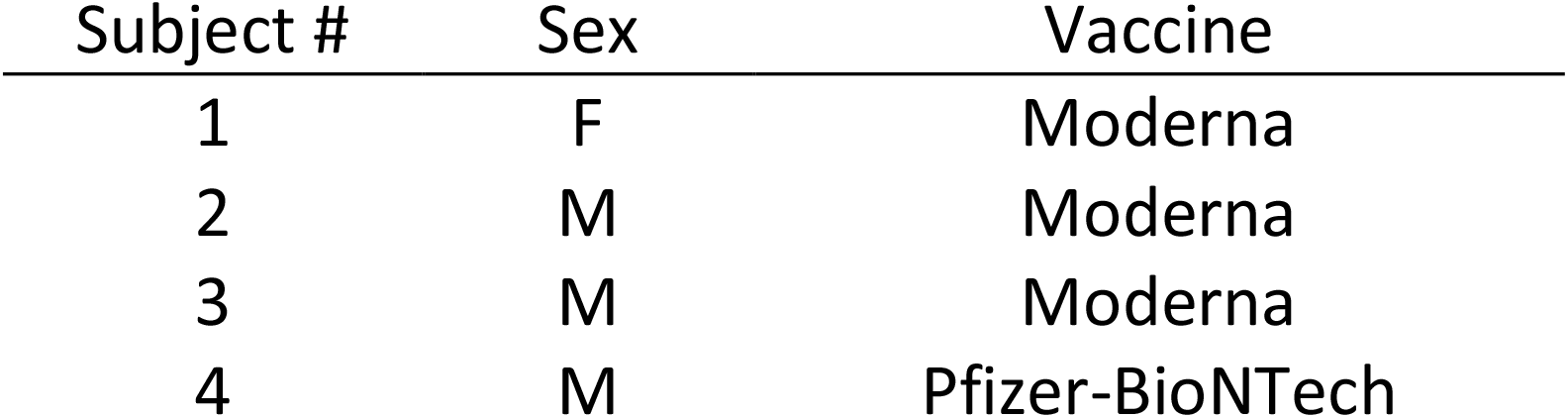
Study subjects that received COVID-19 mRNA vaccine

As shown in Fig. 1, vaccine-induced serum levels of SARS-CoV-2 spike-specific IgG rose exponentially and reached a plateau approximately 18-21 days after the 1^st^ mRNA vaccine dose. After the 2^nd^ vaccine dose, SARS-CoV-2 spike-specific serum IgG increased further, peaking approximately 7 days later, and remaining elevated (78% of peak values) during the additional 20-50 day follow-up period.

**Figure 1.**
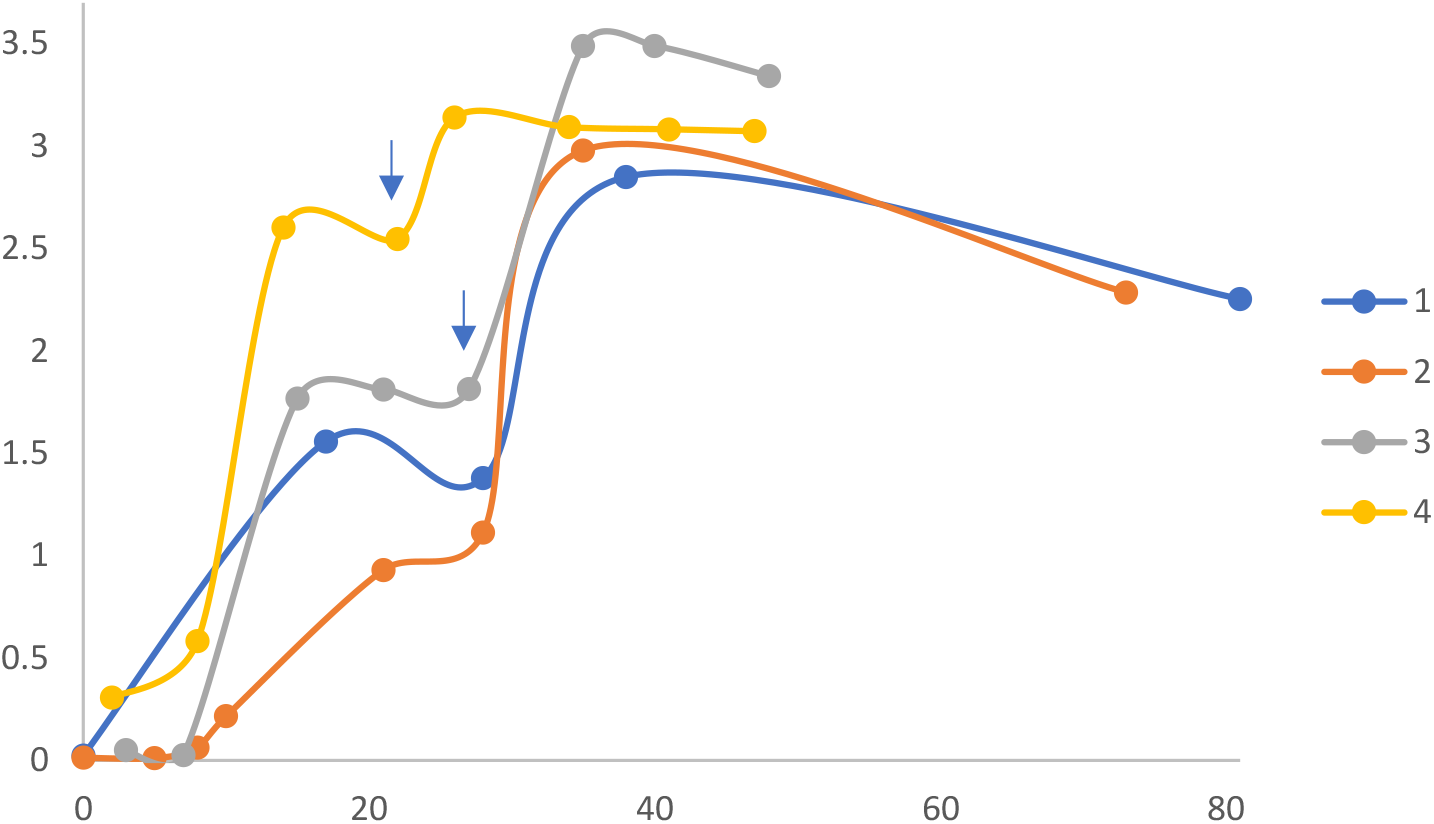
Time course of spike antigen-specific IgG response to COVID-19 mRNA vaccine. ELISAs were performed on serum from 4 subjects described in Table 1, at different time points after vaccination (X-axis, days). Serum IgG levels are proportional to ELISA optical density values (Y-axis). Each symbol represents average ELISA data for a single subject at a single time point. Scatter plots were fitted with a moving average trend line. Note 2^nd^ vaccine dose (blue arrow) was at day 28 for subjects 1-3, and day 21 for subject 4.

COVID-19 mRNA vaccine also elicited spike antigen-specific IgA with similar kinetics of induction and time to maximal levels after the 1^st^ and 2^nd^ vaccine dose (Fig. 2). However, the levels of spike antigen-specific IgA decreased significantly (p<0.001) faster than IgG levels. Spike-specific IgA decreased to an average of 50% peak levels between the 1^st^ and 2^nd^ vaccine shots, and 38% of peak levels within the 50 day follow-up period after the 2^nd^ shot.

**Figure 2.**
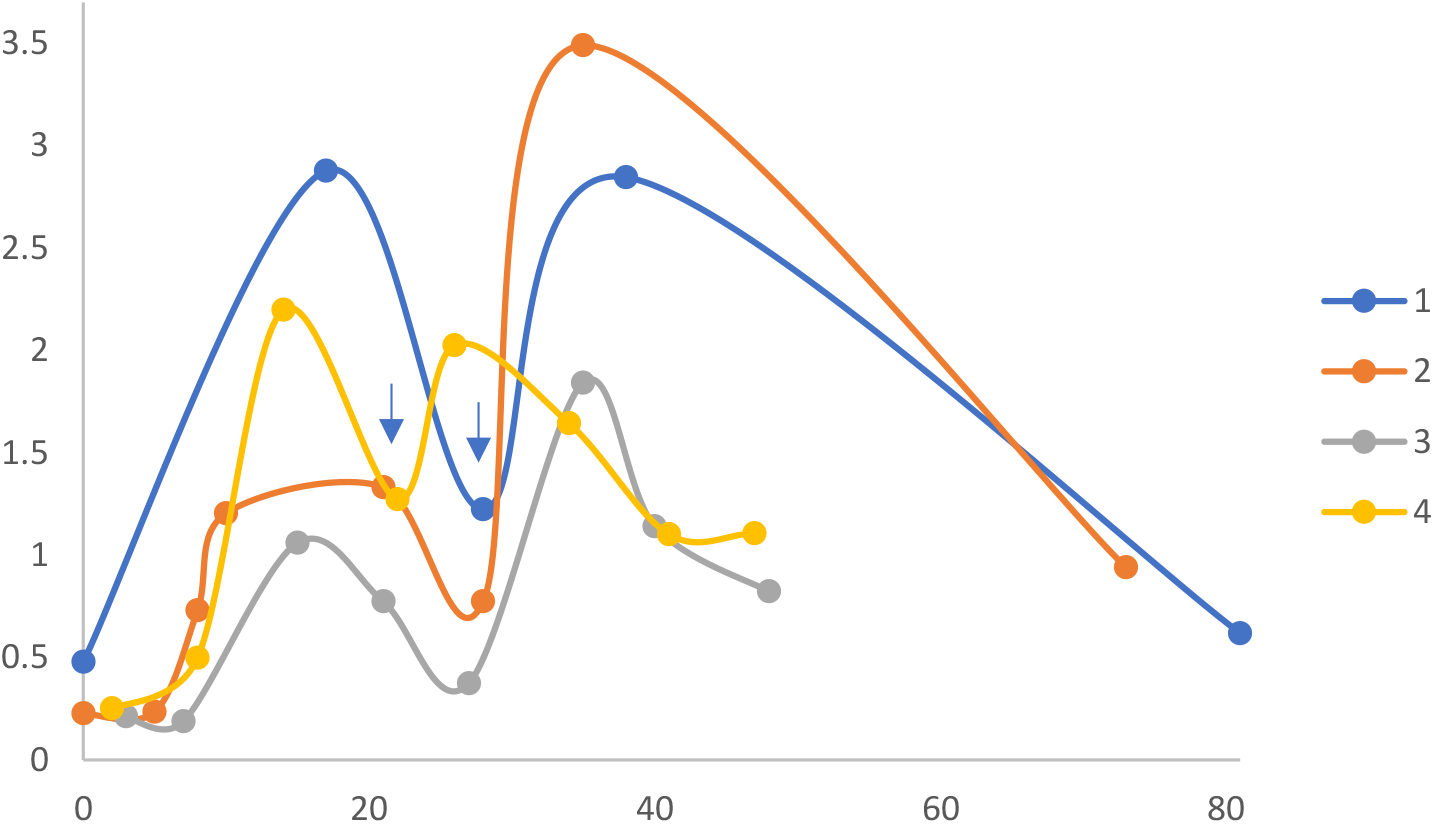
Time course of spike antigen-specific IgA response to COVID-19 mRNA vaccine. ELISAs were performed on serum from 4 subjects described in Table 1, at different time points after vaccination (X-axis, days). Serum IgA levels are proportional to ELISA optical density values (Y-axis). Each symbol represents average ELISA data for a single subject at a single time point. Scatter plots were fitted with a moving average trend line. Note 2^nd^ vaccine dose (blue arrow) was at day 28 for subjects 1-3, and day 21 for subject 4.

The induction and decay of antigen specific IgG and IgA in response to the novel COVID-19 mRNA vaccine are consistent with the known serum ½ lives of the different immunoglobulin isotypes; 21-28 days for gamma and 5-6 days for alpha [17-19]. The rapid decay in serum IgA levels is also consistent with that observed in natural disease among health care workers in a Spanish hospital after 3 months of follow-up [20]. Recent studies document that while SARS-CoV-2 spike-specific serum IgA levels decline quickly after infection, local concentrations at mucosal surfaces persist longer and include dimeric isoforms with potent neutralizing capacity, 15X greater than monomeric IgA [10, 11].

This study focused on serum IgA which has been shown to be clonally related to mucosal IgA, but did not measure vaccine-induced, antigen-specific mucosal IgA levels. Serum IgA may reach mucosal surfaces by transduction or via recirculating IgA secreting plasmablasts with a mucosal homing profile [21-23]. However, “local” B-cells may also undergo isotype class switching in the mucosal microenvironment and secrete IgA with distinct kinetics [24]. The present data underscore the current knowledge gap surrounding COVID-19 mRNA vaccine-induced IgA production and distribution at mucosal sites.

In summary, longitudinal serology of COVID-19 mRNA vaccine recipients highlights important issues related to immunity and monitoring of vaccine responses. The persistence of spike antigen-specific serum IgG following vaccination is hopefully a positive indicator of effective long-lived immunity, and clinical indicator of vaccine responsiveness [25]. In addition to IgG, the data demonstrate COVID-19 mRNA vaccines also elicit antigen-specific IgA, which may be important in preventing transmission as well as infection [26, 27]. Spike-specific serum IgA levels decay significantly faster than spike-specific IgG, however, the “recall” response for both IgG and IgA (time to peak serum levels following the 2^nd^ / booster dose) is significantly shorter than the primary response.

## MATERIALS AND METHODS

### Human subjects

Volunteers from an ongoing serology study of health care workers were recruited to have their SARS-CoV-2 spike antigen-specific antibody levels followed over time after vaccination with SARS-CoV-2 mRNA. Subjects provided 3cc of blood by venipuncture using vacutainer tubes, serum was separated and stored frozen at −80°C until tested by enzyme linked immunosorbent assay (ELISA). The studies were reviewed by the Yale University Human Investigation Committee and ethical approval was given by the Yale university Institutional Review Board.

### ELISA methods

ELISAs were performed as previously described with minor modifications [28, 29]. In short, Triton X-100 and RNase A were added to serum samples at final concentrations of 0.5% and 0.5mg/ml respectively and incubated at room temperature (RT) for 30 minutes before use to reduce risk from any potential virus in serum. 96-well MaxiSorp plates (ThermoFisher, Waltham, MA) were coated with 50 μL/well of recombinant SARS Cov-2 S1 or nucleocapsid protein (Abcam, Cambridge, MA) at a concentration of 1 μg/ml in NaCO_3_ buffer pH 9.6 and incubated overnight at 4°C. The coating buffer was removed, and plates were incubated for 1h at RT with 200 μl of blocking solution (PBS with 3% milk powder). Serum was diluted 1:100 in dilution solution (PBS with 0.05% Tween20, 1% milk powder) and 100 μl of diluted serum was added for one hours at RT. Plates were washed three times with PBS-T (PBS with 0.1% Tween-20) and 50 μL of HRP anti-Human IgG Antibody (Parmingen/BD Biosiences, San Jose, CA) or HRP anti-human IgA (BioLegend, San Diego, CA) were added at 1:2000-fold dilution. After 1 h of incubation at RT, plates were washed three times with PBS-T. Plates were developed with 100 μL of TMB Substrate Reagent Set (BD Biosciences, San Jose, CA) and the reaction was stopped when an internal pooled serum positive control sample reaches an OD of 1.0 at 650 nm, by the addition of 2 N sulfuric acid. Plates were then read at a wavelength of 450 nm with reference wavelength calibration (650nm). Significant differences in rates of decline were calculated based on regression and analysis of variance of geometric means.

## Supporting information

As shown in Fig. 1

## Data Availability

All data will be available in supplemental methods

