## Supplementary material for "Human IgG and IgA responses to COVID-19 mRNA vaccines": As shown in Fig. 1

Table of IgG Data (average values) ……… page 2
Table of IgA Data (average values) ……… page 3

Table of IgG Data
(average values)

| Days Post Vaccine | Subject 1 | Subject 2 | Subject 3 | Subject 4 |
|---|---|---|---|---|
| 0 | | | | |
| 0 | 0.025 | | | |
| 0 | | | | |
| 0 | | 0.014 | | |
| 2 | | | | 0.308 |
| 3 | | | 0.052 | |
| 5 | | 0.011 | | |
| 7 | | | | |
| 7 | | | 0.026 | |
| 8 | | 0.062 | | |
| 8 | | | | 0.584 |
| 10 | | 0.216 | | |
| 14 | | | | |
| 14 | | | | 2.603 |
| 15 | | | 1.769 | |
| 17 | 1.558 | | | |
| 21 | | 0.93 | | |
| 21 | | | 1.812 | |
| 22 | | | | |
| 22 | | | | 2.547 |
| 26 | | | | 3.141 |
| 27 | | | 1.813 | |
| 28 | 1.379 | | | |
| 28 | | 1.113 | | |
| 34 | | | | 3.093 |
| 35 | | 2.98 | | |
| 35 | | | 3.49 | |
| 38 | 2.85 | | | |
| 40 | | | 3.49 | |
| 41 | | | | 3.083 |
| 43 | | | | |
| 44 | | | | |
| 47 | | | | 3.073 |
| 48 | | | 3.343 | |
| 58 | | | | |
| 65 | | | | |
| 73 | | 2.286 | | |
| 81 | 2.254 | | | |

Table of IgA data (Average values)

| Days Post Vaccine | Subject 1 | Subject 2 | Subject 3 | Subject 4 |
|---|---|---|---|---|
| 0 | | | | |
| 14 | | | | |
| 43 | | | | |
| 65 | | | | |
| 0 | 0.479 | | | |
| 17 | 2.878 | | | |
| 28 | 1.224 | | | |
| 38 | 2.846 | | | |
| 81 | 0.619 | | | |
| 0 | | | | |
| 7 | | | | |
| 22 | | | | |
| 44 | | | | |
| 58 | | | | |
| 0 | | 0.228 | | |
| 5 | | 0.235 | | |
| 8 | | 0.731 | | |
| 10 | | 1.205 | | |
| 21 | | 1.333 | | |
| 28 | | 0.774 | | |
| 35 | | 3.49 | | |
| 73 | | 0.941 | | |
| 3 | | | 0.213 | |
| 7 | | | 0.188 | |
| 15 | | | 1.06 | |
| 21 | | | 0.774 | |
| 27 | | | 0.375 | |
| 35 | | | 1.843 | |
| 40 | | | 1.142 | |
| 48 | | | 0.823 | |
| 2 | | | | 0.252 |
| 8 | | | | 0.499 |
| 14 | | | | 2.198 |
| 22 | | | | 1.274 |
| 26 | | | | 2.026 |
| 34 | | | | 1.645 |
| 41 | | | | 1.102 |
| 47 | | | | 1.107 |
